# Optimal waist circumference cut-off points for predicting Metabolic Syndrome among females of reproductive age in Wakiso district, central Uganda

**DOI:** 10.1101/2024.03.08.24303971

**Authors:** David Lubogo, Henry Wamani, Roy William Mayega, Christopher Garimoi Orach

## Abstract

**Background:** Metabolic Syndrome (MetS) poses a significant challenge to global public health, due to its strong association with Type 2 diabetes and cardiovascular disease. Waist circumference (WC) is a convenient metric for diagnosing MetS. Our study sought to establish waist circumference cut-offs that predict MetS in females of reproductive age residing in Wakiso district in central Uganda.

**Methods:** The data collected were from a cross-sectional study conducted in Wakiso district, central Uganda, involving 697 randomly selected females aged 15 to 49 between June 9^th^ and August 17^th,^ 2021. Data included MetS components: WC, High-Density Lipoprotein (HDL) Cholesterol, triglycerides, blood pressure, and fasting blood sugar. MetS status was identified based on two or more MetS components excluding WC. ROC analysis established the average optimal WC. The accuracy and performance of the cut-off points were evaluated through sensitivity, specificity, Positive likelihood ratio, and the Youden index.

**Results:** Of the 697 participants, 49.9% had two or more MetS risk factors. For females aged 15-49 years, the average optimal WC cut-off was 80.3 cm. Variations in optimal WC thresholds were observed across different age groups: 97.4 cm for (15-24 years), 79.9 cm (25-34 years), 85.6 cm (35-44 years), and 91.1 cm (45-54 years) respectively. The area under the ROC curve for these age groups ranged from 0.78 to 0.86, indicating good discriminatory capability. The sensitivity ranged from 85% to 97%, specificity from 58% to 88%, and the Youden Index from 0.557 to 0.729.

**Conclusions:** A waist circumference of 80.3 cm is the optimal threshold for identifying Metabolic Syndrome in females between the ages of 15 and 49 years in the setting. This finding concurs with the guidance set forth by the International Diabetes Federation. Additionally, study participants had varying WC cut-offs ranging from 79.4 cm to 91.1 cm, depending on their age.

## Introduction

Metabolic Syndrome (MetS) refers to the accumulation of various risk factors related to cardiac and metabolic health (1, 2). Among these risk factors is the presence of abdominal obesity, low HDL Cholesterol, elevated triglycerides, high blood pressure, and elevated fasting blood sugar, (3, 4). MetS is a significant global public health challenge, contributing to high morbidity and mortality, additionally, its occurrence is growing in Africa (5–8). The failure to tackle MetS greatly raises the chances of developing Type 2 diabetes and cardiovascular disease (9–11). MetS is more prevalent among females than males (6, 9), directly affects their reproductive role, and contributes to adverse perinatal (12, 13) and maternal outcomes(14).

The measurement of waist circumference (WC) serves as a simple and practical indicator for assessing abdominal obesity, which is a key component of Metabolic Syndrome (15, 16). The World Health Organization (WHO) and other scientific organizations strongly recommend using waist circumference measurement in screening, diagnosis, and routine clinical care to guide patient management (4, 17). This requires an acceptable waist circumference cut-off for risk stratification. However, establishing universally accepted WC cut-offs has been challenging due to differences in abdominal tissue composition among ethnic groups (18). This is because genetics, environmental, and lifestyle elements could impact abdominal obesity (19) and overall body composition (20). In the interim, Uganda and other countries in sub-Saharan Africa use WC cut-offs endorsed by international scientific organizations including the WHO (3, 20). Nevertheless, it is still unclear whether established thresholds, such as the ≥ 80 cm suggested for females, are valid and appropriate for the African population. Hence, it the necessary to conduct research on the diverse African population groupings including Uganda and to establish sex-specific WC cut-offs as recommended by WHO and others (4, 20).

To date, most studies on optimal waist circumference cut-off points have been conducted in high-income countries. In low-income countries such as Uganda, the optimal ethnic WC cut-off has not been established. Therefore, this study aimed at determining the average waist circumference and age-specific cut-off points that are optimal for predicting MetS among females of reproductive age in Wakiso district, central Uganda. The hypothesis for the study was that the optimal waist circumference for females aged 15-49 years might not be equal to 80.0cm.

## Methods

### Study design

We carried out a cross-section study in Wakiso district between June 9, 2021 and August 17, 2021. Our study involved a random sample of female participants from 15 to 49 years of age.

### Study population

The study population included females aged 15-49 years residing in Wakiso district, central Uganda. Participants were randomly selected from urban and rural communities to ensure diversity. Eligible participants had resided in Wakiso for at least a year and were identified from a database of 800 individuals, who had anthropometric, physical, and biochemical tests carried out on them to determine the prevalence of MetS in the district.

We excluded participants who were pregnant, those within six weeks after childbirth, alcoholics, smokers, those with chronic diseases such as diabetes mellitus, hypertension, hyperlipidaemia, allergic disorders, depression, and those with known carrier states of hepatitis B, hepatitis C, or human Immunodeficiency virus.

### Study setting

The study was conducted in Wakiso district, situated in central Uganda, with an estimated total population of 3,105,700, based on the 2014 census. More than half (52.7%) of the residents are females. Of these, 793,770 are in the reproductive age bracket of 15-49 years (21). The district comprises two counties, namely Kyaddondo and Busiro, and four Municipal Councils - Entebbe, Nansana, Kira, and Makindye-Ssabagabo that together constitute eight sub-counties, eight town councils, 148 parishes, and 704 villages. The Uganda Bureau of Statistics provided the sampling frame for the study and it included 32 randomly selected enumeration areas in the district. Twenty-eight (28) households were sampled per enumeration area.

### Sample size calculation

The sample size was computed using the OpenEpi, Version 3.01, open-source calculator for a proportion or descriptive study (22). Using a population size for a finite population of 1,000,000, confidence limits of 5%, a design effect of 2.0., and a prevalence (p) of 40.2% +/-5 obtained from a study conducted among females in an urban population in Kenya to determine MetS prevalence (23).

A sample size of 739 was obtained; after adjusting for a response rate of 90%, the sample size was 813. The final sample used in the study was 800 respondents. We estimated a MetS prevalence of 15% and expected 120 respondents to be with MetS out of the total sample size of 800. This sample was considered appropriate for assessing the sensitivity and specificity of WC as a predictor for MetS in this sample.

### Measurements and variables

We used the population sample data set from a prevalence study to obtain participant data on anthropometric measurements (waist circumference), physical measurements (blood pressure), and biochemical measurements (fasting plasma glucose, triglycerides, and low HDL Cholesterol) collected as per the WHO STEP wise approach (24).

Quality control measures were undertaken to ensure the data for the prevalence study were of quality. They included the employment of well-trained data collectors/research assistants (female nurses and graduate nutritionists) who were knowledgeable of the study protocol and had skills in using the Open Data Kit (ODK) for data collection. The data collection tools were field tested from a district near the study district. The data collected in the field were entered in the ODK tool embedded with skip patterns and constraints to ensure data quality and reduce bias. Quantitative data (ODK data) was downloaded from the server and exported to Stata version 14 for cleaning. This ensured reasonable quality control and data management for this study.

### Study variables

#### Dependent variable

Metabolic Syndrome was assessed by determining if an individual had three or more of the five components: low HDL cholesterol or treatment for it, high triglycerides or treatment for it, elevated fasting blood sugar or diabetes diagnosis, or diabetes treatment; high blood pressure or treatment for it, and increased waist circumference, as defined in the Joint Interim Statement (JIS) (4)

#### Data analysis

Data were analysed using Stata statistical software (SE/14.0, StatCorp, College Station, TX, USA (25). Descriptive statistics were employed to summarise participant characteristics. Data on continuous variables were summarized using the independent sample T-test and categorical variables were analyzed using the Chi-square test. The results were presented as means with standard deviation and frequencies/percentages, respectively.

Waist circumference cut-off points for various age groups (15-45, 15-24, 25-34, 35-44, 45-54 years) were evaluated using sensitivity and specificity techniques to identify the most effective predictive values for MetS within the study age groups.

The research investigated the Area under the Curve (AUC) for different components of MetS such as elevated fasting blood sugar, high triglycerides, high blood pressure and low HDL cholesterol. To conduct this analysis, an ROC curve was plotted for the association between waist circumference (WC) and two or more components of MetS.

The optimal cut-off points for WC, which achieves maximum accuracy, were determined by plotting sensitivity and (1-specificity) on a graph.

The study employed the Youden index method (26) expressed as [Maximum (sensitivity + specificity - 1)] to ascertain the optimal WC values and also calculated the positive predictive values (PPV) and negative predictive values (NPV).

#### Ethical considerations

We obtained approval from the Higher Degrees Research and Ethics Committee of the School of Public Health, College of Health Sciences, Makerere University (Protocal number: MakSPH-REC, 071) and the National Council for Science and Technology of Uganda (Registration number: UNCST, HS1281ES). All study participants provided written informed consent and were assured of the confidentiality of their information.

## Results

### Flow of participants

The figure in S1 Fig shows the flow diagram of the study participants

**S1 Fig:** Flow diagram of study participants

### Clinical and biochemical characteristics of the study participants

The study involved 697 women of reproductive age. Participants with incomplete data were not included in the analysis (S6 Fig). On average, they were 29.7 years old (± 9.07), with a mean waist circumference of 86.4 cm (± 13.8) and the mean body mass index (BMI) of 26.9 (±5.8). Almost half of the participants (49.9%) exhibited two or more risk factors linked to MetS (≥ 2 MetS factors), as indicated in Table 1.

**Table 1:** Clinical and biochemical characteristics of study participants.

| Variables | Total |
| --- | --- |
| | Mean ± SD |
| Number of participants (%) | 697 (100%) |
| Age (years) | 29.7 ± 9.07 |
| Multiple risk factor aggregation (MRFA) ($\geq 2$ MetS factors) | 49.93% |
| Triglyceride (mmol/l) | 1.13 ± 0.62 |
| HDL cholesterol (mmol/l) | 1.33 ± 0.38 |
| Fasting blood sugar (mmol/l) | 4.56 ± 1.16 |
| Body mass index (kg/m<sup>2</sup>) | 26.9 ± 5.8 |
| Systolic BP (mmHg) | 117.6 ± 15.3 |
| Diastolic BP (mmHg) | 76.3 ± 11.2 |
| Waist circumference (cm) | 86.4 ± 13.8 |

### Age-specific evaluation of optimal thresholds for waist circumference using different classification metrics

For the age group 15-45 years, the optimal threshold was 80.3 cm. The Youden index, specificity and sensitivity were 0.604, 67% and 94% respectively, and the AUC curve was 0.848 (95% CI 0.819 - 0.877).

The optimal WC cut-off points varied among the various age groups, ranging from 79.4 cm for females aged 15 to 24 years to 91.1 cm for those aged 45 to 54 years. Across different age groups, the area under the ROC curve values fluctuated between 0.78 and 0.86, with the 45-54-year-old age group showing the highest AUC of 0.891. The 45-54 years age group showed the highest discriminative power (AUC =0.891), with specificity (88 %), Youden index (0.729), and a likelihood ratio of (7.08), as shown in Table 2.

**Table 2:** Age - stratified Waist circumference cut-off points and their classification metrics for MetS risk.

| Variables | Age group |  |  |  |  |
| --- | --- | --- | --- | --- | --- |
|  | 15-24 years | 25-34 years | 35-44 years | 45-54 years | 15-45 years |
| Empirical optimal WC cut-off point (cm) | 79.4 | 79.9 | 85.6 | 91.1 | 80.3 |
| Area under the ROC curve at the cut point | 0.83 | 0.78 | 0.80 | 0.86 | 0.80 |
| Area under ROC curve (95% CI) | 0.8553(0.803 - 0.907) | 0.804(0.750 - 0.858) | 0.852(0.786 - 0.918) | 0.891 (0.790 - 0.993) | 0.848 (0.819 - 0.877) |
| Sensitivity at cut point (%) | 90 | 97 | 85 | 85 | 94 |
| Specificity at cut point (%) | 77 | 58 | 75 | 88 | 67 |
| Youden index (J) | 0.665 | 0.557 | 0.604 | 0.729 | 0.604 |
| LR+ = sensitivity / (1 - specificity) | 3.913 | 2.3095 | 3.4 | 7.08 | 2.85 |
| Presence of less than 2 metabolic syndrome risk factors (no.) | 137 | 134 | 60 | 17 | 348 |
| Presence of 2 or more metabolic syndrome risk factors (no.) | 89 | 118 | 103 | 39 | 349 |
| **Total** | **226** | **252** | **163** | **56** | **697** |

### ROC curve analysis, for waist circumference in reproductive-age women in Wakiso district, for detecting MetS risk factors per age group

Figures in S1-S5 Figs show the ROC curves for WC to predict the presence of 2 or more MetS risk factors among females of reproductive age for the various age groups;

**S2 Fig:** ROC curve for 15-45 years old females in Wakiso district, 2023

**S3 Fig:** ROC curve for 15-24 years old females in Wakiso district, 2023

**S4 Fig:** ROC curve for 25-34 years old females in Wakiso district, 2023

**S5 Fig:** ROC curve for 35-44 years old females in Wakiso district, 2023

**S6 Fig:** ROC curve for 45-54 years old females in Wakiso district, 2023

## Discussion

The rise in the global prevalence of MetS calls for more research towards early and accurate diagnosis of MetS to provide preventive measures and timely intervention for MetS.

Our study revealed that the mean WC cut-off point of 80.3 cm accurately predicted MetS in females aged 15-49 years in an urban population in Uganda. However, optimal waist circumference varied by age groups (15-24 years, 25-34 years, 35-44 years, and 45-54 years) being 79.4 cm, 79.9 cm, 85.6 cm, and 91.1 cm respectively. This study is the first to use the Joint Interim Statement to establish these cut-off thresholds in Uganda.

Our study has identified an optimal WC threshold value that closely approximates the recommended cut-off of 80.0 cm as specified by the International Diabetes Federation for diagnosing Metabolic Syndrome (MetS)(27). Similar results have been found in prior studies conducted across Africa and other locations. An optimal WC of 80.5 cm was found among female university employees in Angola(28). Elsewhere, some studies among a multiethnic Malaysian population (29) and in a Japanese population (30) found that 80.0 cm was an optimal WC for women. The consistency of our findings with those of other researchers strengthens the validity of our findings and reinforces the global relevance of the established diagnostic criteria. From a public health perspective, policymakers and healthcare professionals may utilize these findings to implement cost-effective and efficient screening programs to enable the early detection and management of MetS.

Our findings, however, differed from a study in Ethiopia, which revealed the optimal WC at 78.0 cm (31). Higher optimal WC cut off points of ≥ 82.3 cm and 92.0 cm were found in studies conducted among women in Botswana and South Africa, respectively(15, 32). These variations underscore the importance of considering region-specific factors when considering WC cutoff points for assessing women’s health. The observed variation might be attributed to differences in factors such as genetics and body composition across different geographical regions. For instance, countries with higher prevalences of obesity, such as South Africa, often have higher optimal WC cut-off points (15, 33).

This study also revealed varying age-specific WC thresholds for various age groups: 79.4 cm for people aged 15-24 years, 79.9 cm for those aged 25-34 years, 85.6 cm for the 35-44 age group, and 91.1 cm for those aged 45-54 years respectively. The age-specific cut-off points highlight the variability of MetS risk across various age groups. The younger individuals showed risk factors even with slightly lower waist circumference levels. This emphasizes the importance of early detection and timely intervention in this group. Conversely, elderly individuals exhibited higher waist circumference cut-off points. This is in agreement with other studies (34). Probable reasons for this could be due to changes in body composition with age resulting in reduced muscle mass and again in body fat, and a reduction in metabolic rate with age resulting in weight gain if dietary intake and physical activity remained constant.

Therefore, to effectively combat the rising burden of cardiovascular diseases, healthcare providers and policymakers should include age-specific thresholds for waist circumference in screening and intervention protocols for Metabolic syndrome.

### Diagnostic accuracy of the waist circumference cut-off points to detect MetS risk factors

Different metrics were employed to assess the precision of detecting Metabolic Syndrome, including AUC, PPV, specificity and sensitivity, with reference to a specific waist circumference threshold.

When we measured the performance of the AUC values within the 15-24 age range, the AUC reached 0.83. This data suggests that the WC cut-off point can serve as a reasonably accurate indicator of MetS risk in individuals within this age range. For the 25-34 years age group, the AUC was 0.78, this demonstrates moderate performance. In the 35-44 years age category, the AUC was 0.80, this suggests a relatively good ability to distinguish those at risk for MetS. In the 45-54 years age group, the AUC was 0.86., this age group exhibited the highest AUC values, and thus the most robust discriminative power. This suggests that this group may be the most appropriate target for MetS screening using WC. Considering all individuals aged 15-45 years, the overall AUC was 0.80, indicating good performance in identifying MetS. Normally, an AUC of 1 indicates a perfect classifier with the model yielding a 100% true positive rate and 0% false positive rate across all possible threshold values. An AUC of 0 shows that the classifier performs no better than chance, is not able to distinguish between two classes and yields a 0% true positive rate and 100% false positive rate (35). Most classifiers aim for an AUC above 0.5, and that indicates predictive power.

These findings hold important implications for healthcare professionals and researchers, as they provide valuable guidance for developing protocols for screening MetS that are specific to different age groups.

### Effectiveness of waist circumference cut-off points in terms of sensitivity and specificity

Considering sensitivity and specificity values, in the 15-24 age group, a WC of 79.4 cm was obtained with 90% sensitivity and 77% specificity. The high sensitivity suggests that it is effective in identifying Metabolic Syndrome. However, the moderate specificity indicates some potential for false positives.

The 25-34 age group showed a WC cut-off of 79.9 cm and had the highest sensitivity of 97%. This means that nearly all individuals with Metabolic Syndrome in this age group are accurately identified. However, a lower 58% specificity may imply concerns about potential false positives, and thus there is a need to do confirmation tests.

The 35-44 age category, had an 85.6 cm cut-off, and balanced sensitivity (85%) and specificity (75%). This implies that it is reasonably practical in identifying those with Metabolic Syndrome and can minimize the risk of false positives, making it a valuable tool for screening.

In the 45-54 age group, there was a 91.1 cm cut-off point, with an 85% sensitivity and 88% specificity. This implies that this cut-off can accurately identify individuals at risk and can minimise false positives. For the age range of 15-45 years, we obtained an 80.25 cm cut-off point, with 94% sensitivity and 67% specificity. This shows that has the potential to serve as an efficient screening tool. However, the lower specificity of 67% calls for further evaluation to confirm diagnoses.

Generally, these age-specific WC cut-off points can be used to offer tailored strategies for MetS risk assessment and underscore the importance of age in risk prediction and patient management. Further research may be needed to validate and expand these findings for broader clinical and public health applications.

### Youden Index and Positive Likelihood Ratios (LR+)

Our study evaluated age-specific waist circumference (WC) cut-off points and their associated Youden index (J) and Positive Likelihood Ratios (LR+) to determine the diagnostic performance across different age groups.

Our study identified distinctions among the different age categories. The 45-54-year-old age group showed the highest Youden index (J) of 0.729, indicating a higher overall accuracy than the other age groups. A value of one (1) indicates perfect accuracy, and the closer J is to 1, the better the test’s overall performance (36). Furthermore, it showed the highest LR+ value of

### 7.08. This implies that among individuals within this age group, a positive test result is about

7.08 times more likely to occur in individuals with Metabolic Syndrome compared to those without the syndrome. This makes it a robust diagnostic tool. The 25-34 years age group showed a slightly lower Youden index (J) of 0.557 (indicating slightly lower overall accuracy) compared to the 15-24 years group with a Youden index (J) of 0.665. The 25-34 years age group showed an LR+ of 2.3095, demonstrating valuable diagnostic utility. A LR+ of 2.3095 suggests that individuals diagnosed with Metabolic Syndrome have a waist circumference above the cut-off point approximately 2.3 times more often than those without Metabolic Syndrome. Although this LR+ was slightly lower than the 15-24 years group LR+ (Likelihood Ratio Positive) of 3.913, this LR+ is still a valuable indicator of diagnostic strength. The 35-44 years and the broader 15-45 years age range both demonstrated intermediate Youden index (J) values of 0.604, with corresponding LR+ values of 3.4 and 2.85, respectively.

The study demonstrates that for urban Ugandan females aged 15-49 years, 80.3 cm waist circumference is the mean optimal threshold for MetS prediction. Furthermore, waist circumference cut-off points varied depending on age group as indicated; 79.4 cm (15-24 years), 79.9 cm (25-34 years), 85.6 cm (35-44 years), and 91.1 cm (45-54 years). Our findings provide useful information for healthcare professionals when considering age-tailored risk assessment strategies for Metabolic Syndrome to enhance the clinical relevance of WC measurements.

### Study limitations

We used a cross-sectional study design. MetS is a condition that develops over time, and the measurement of WC in a single time point may not fully capture its dynamic nature. A longitudinal study would be more appropriate for confirming the temporal relationships observed in the study findings.

Our study focused exclusively on females of reproductive age in Wakiso district, central Uganda. Consequently, while the findings are relevant to women in a similar setting, the results may not be generalizable to the entire Ugandan population. Conducting nationally representative studies on optimal waist circumference in the future would help address this limitation and provide a better understanding of MetS in Uganda.

## Conclusions and recommendations

We found that a waist circumference of 80.3 cm was the mean optimal threshold for identifying Metabolic Syndrome in females between the ages of 15 and 49 years in Wakiso district, central Uganda. Furthermore, participants exhibited waist circumference cut-offs ranging from 79.4 cm to 91.1 cm based on age group. This study highlights the significance and precision of the existing waist circumference guidelines in healthcare. This study calls for the utilization of a waist circumference cut-off threshold to predict Metabolic Syndrome among reproductive-age females in Uganda, based on age group. The findings of this research may be relevant to healthcare providers, policymakers, public health practitioners and researchers. Further research in a wider range of populations encompassing both rural and urban settings is needed to validate these waist circumference cut-off points.

## Data Availability

Data already provided as part of the submitted article.

## Acknowledgements

We appreciate all the study participants and research assistants who supported the data collection process. We thank all the local leaders and the district health management team members in Wakiso district who facilitated the research teams. We appreciate Dr Arthur Bagonza (PhD) and Dr Freddie Kitutu (PhD) for reviewing the manuscript.

## Supporting Information

**S1 Fig:** ROC curve for 15-45 years old females in Wakiso district, 2023

**S2 Fig:** ROC curve for 15-24 years old females in Wakiso district, 2023

**S3 Fig:** ROC curve for 25-34 years old females in Wakiso district, 2023

**S4 Fig:** ROC curve for 35-44 years old females in Wakiso district, 2023

**S5 Fig:** ROC curve for 45-54 years old females in Wakiso district, 2023

## References

1. Bovolini A, Garcia J, Andrade MA, Duarte JA. Metabolic syndrome pathophysiology and predisposing factors. International Journal of Sports Medicine. 2021;42(03):199–214.

2. Wang HH, Lee DK, Liu M, Portincasa P, Wang DQ-H. Novel insights into the pathogenesis and management of the metabolic syndrome. Pediatric gastroenterology, hepatology & nutrition. 2020;23(3):189.

3. Alberti KGMM, Zimmet P, Shaw J. Metabolic Syndrome—a new world-wide definition. A consensus statement from the International Diabetes Federation. Diabetic medicine. 2006;23(5):469–80.

4. Alberti K, Eckel RH, Grundy SM, Zimmet PZ, Cleeman JI, Donato KA, et al. Harmonising the Metabolic Syndrome a Joint Interim Statement of the International Diabetes Federation Task Force on Epidemiology and Prevention; National Heart, Lung, and Blood Institute; American Heart Association; World Heart Federation; International Atherosclerosis Society; and International Association for the study of Obesity. Circulation. 2009;120(16):1640–5.

5. Okafor CI. The metabolic syndrome in Africa: Current trends. Indian journal of endocrinology and metabolism. 2012;16(1):56–66.

6. Bowo-Ngandji A, Kenmoe S, Ebogo-Belobo JT, Kenfack-Momo R, Takuissu GR, Kengne-Ndé C, et al. Prevalence of the metabolic syndrome in African populations: A systematic review and meta-analysis. PLoS One. 2023;18(7):e0289155.

7. Saklayen MG. The global epidemic of the metabolic syndrome. Current hypertension reports. 2018;20(2):1–8.

8. Noubiap JJ, Nansseu JR, Lontchi-Yimagou E, Nkeck JR, Nyaga UF, Ngouo AT, et al. Geographic distribution of metabolic syndrome and its components in the general adult population: A meta-analysis of global data from 28 million individuals. Diabetes research and clinical practice. 2022;188:109924.

9. Faijer-Westerink HJ, Kengne AP, Meeks KA, Agyemang C. Prevalence of metabolic syndrome in sub-Saharan Africa: A systematic review and meta-analysis. Nutrition, Metabolism and Cardiovascular Diseases. 2020;30(4):547–65.

10. Hwang Y-C, Jee J-H, Oh EY, Choi Y-H, Lee M-S, Kim K-W, et al. Metabolic syndrome as a predictor of cardiovascular diseases and type 2 diabetes in Koreans. International journal of cardiology. 2009;134(3):313–21.

11. Mohamed SM, Shalaby MA, El-Shiekh RA, El-Banna HA, Emam SR, Bakr AF. Metabolic syndrome: risk factors, diagnosis, pathogenesis, and management with natural approaches. Food Chemistry Advances. 2023;3:100335.

12. dos Prazeres Tavares H, Dos Santos DCDM, Abbade JF, Negrato CA, De Campos PA, Calderon IMP, et al. Prevalence of metabolic syndrome in non-diabetic, pregnant Angolan women according to four diagnostic criteria and its effects on adverse perinatal outcomes. Diabetology & Metabolic Syndrome. 2016;8:1–12.

13. Bo S, Menato G, Gallo M-L, Bardelli C, Lezo A, Signorile A, et al. Mild gestational hyperglycemia, the metabolic syndrome and adverse neonatal outcomes. Acta obstetricia et gynecologica Scandinavica. 2004;83(4):335–40.

14. Grieger JA, Bianco-Miotto T, Grzeskowiak LE, Leemaqz SY, Poston L, McCowan LM, et al. Metabolic syndrome in pregnancy and risk for adverse pregnancy outcomes: A prospective cohort of nulliparous women. PLoS medicine. 2018;15(12):e1002710.

15. Crowther NJ, Norris SA. The current waist circumference cut point used for the diagnosis of metabolic syndrome in sub-Saharan African women is not appropriate. PloS one. 2012;7(11):e48883.

16. Fang H, Berg E, Cheng X, Shen W. How to best assess abdominal obesity. Current opinion in clinical nutrition and metabolic care. 2018;21(5):360.

17. Ross R, Neeland IJ, Yamashita S, Shai I, Seidell J, Magni P, et al. Waist circumference as a vital sign in clinical practice: a Consensus Statement from the IAS and ICCR Working Group on Visceral Obesity. 2020. Report No.: 1759-5029 Contract No.: 3.

18. Misra A, Wasir JS, Vikram NK. Waist circumference criteria for the diagnosis of abdominal obesity are not applicable uniformly to all populations and ethnic groups. Nutrition. 2005;21(9):969–76.

19. Nicolaidis S. Environment and obesity. Metabolism. 2019;100:153942.

20. WHO. Waist circumference and waist-hip ratio: report of a WHO expert consultation, Geneva, 8–11 December 2008. Geneva; 2011.

21. UBOS. Population and Censuses Kampala: UBOS; 2020 [Available from: https://www.ubos.org/explore-statistics/20/.

22. Sullivan K, Dean A. OPENEPI: A Web-based epidemiologic and statistical calculator for Public Health. 2009; 124: 471–474. 2009.

23. Kaduka LU, Kombe Y, Kenya E, Kuria E, Bore JK, Bukania ZN, et al. Prevalence of metabolic syndrome among an urban population in Kenya. Diabetes Care. 2012;35(4):887–93.

24. WHO. WHO STEPS surveillance manual: the WHO STEPwise approach to chronic disease risk factor surveillance. Geneva: World Health Organization; 2005. Report No.: 9241593830.

25. StataCorp L. StataCorp stata statistical software: Release 14. StataCorp LP: College Station, TX, USA. 14.1 ed2015.

26. Fluss R, Faraggi D, Reiser B. Estimation of the Youden Index and its associated cutoff point. Biometrical Journal: Journal of Mathematical Methods in Biosciences. 2005;47(4):458–72.

27. Alberti, Kurt George Matthew Mayer, Zimmet Paul, Shaw. J. Metabolic Syndrome—a new world-wide definition. A consensus statement from the International Diabetes Federation. Diabetic medicine. 2006;23(5):469–80.

28. Magalhaes P, Capingana DP, Mill JG. Prevalence of the metabolic syndrome and determination of optimal cut-off values of waist circumference in university employees from Angola. South African Journal of Diabetes and Vascular Disease. 2015;12(1):35–40.

29. Cheong KC, Ghazali SM, Hock LK, Yusoff AF, Selvarajah S, Haniff J, et al. Optimal waist circumference cut-off values for predicting cardiovascular risk factors in a multi-ethnic Malaysian population. Obesity research & clinical practice. 2014;8(2):e154–e62.

30. Ogawa D, Kahara K, Shigematsu T, Fujii S, Hayakawa N, Okazaki M, et al. Optimal cut-off point of waist circumference for the diagnosis of metabolic syndrome in Japanese subjects. Journal of Diabetes Investigation. 2010;1(3):117–20.

31. Sinaga M, Worku M, Yemane T, Tegene E, Wakayo T, Girma T, et al. Optimal cut-off for obesity and markers of metabolic syndrome for Ethiopian adults. Nutrition journal. 2018;17(1):1–12.

32. Tladi D, Mokgatlhe L, Shaibu S, Nell T, Mitchell R, Mokgothu C, et al. Determination of optimal cut-off values for waist circumferences used for the diagnosis of the metabolic syndrome among Batswana adults (ELS 32). Cardiovascular Journal of Africa. 2020;31(6):314–8.

33. Motala AA, Esterhuizen T, Pirie FJ, Omar MA. The prevalence of metabolic syndrome and determination of the optimal waist circumference cutoff points in a rural South African community. Diabetes care. 2011;34(4):1032–7.

34. Subramoney S, BjöRkeluND C, Guo X, Skoog I, Bosaeus I, Lissner L. Age-related differences in recommended anthropometric cut-off point validity to identify cardiovascular risk factors in ostensibly healthy women. Scandinavian journal of public health. 2014;42(8):827–33.

35. Kohn MA, Newman TB. The walking man approach to interpreting the receiver operating characteristic curve and area under the receiver operating characteristic curve. Journal of Clinical Epidemiology. 2023;162:182–6.

36. Shan G. Improved confidence intervals for the Youden index. PloS one. 2015;10(7):e0127272.

